# General-Purpose vs. Domain-Specific Large Language Models in Antibiotic Clinical Decision-Making: A Double-Blind Evaluation with a 2×2 Factorial Design

**DOI:** 10.64898/2026.07.11.26357814

**Authors:** Yang Liu, Changjing Zhang, Feifei Wang, Wei Xu, Yunhe Zhang, Shaolin Ma, Haitao Zhang

**Affiliations:** Department of Critical Care Medicine, Shanghai East Hospital, Tongji University School of Medicine, Shanghai, China

**Keywords:** large language models, antimicrobial stewardship, clinical decision support, chain-of-thought prompting, prompt engineering, artificial intelligence

## Abstract

**Background:** Antimicrobial resistance poses a major threat to global public health. Large language models (LLMs) offer new possibilities for optimizing antibiotic prescribing decisions, but the capabilities of general-purpose versus domain-specific medical LLMs under different prompting strategies remain to be clarified.

**Methods:** This double-blind, randomized-sequence evaluation used a 2×2 factorial design comparing four AI conditions—the domain-specific model MedGo and the general-purpose model DeepSeek V3.5, each under standard direct prompting and chain-of-thought (CoT) prompting—alongside real physician prescriptions across 59 complex inpatient infection cases. Five parallel regimens were generated per case and independently evaluated by three senior clinicians (1–5 comprehensive score and five domain sub-scores). ChatGPT 5.2 was additionally assessed as an automated evaluation tool.

**Results:** Score ranking: real physicians (4.76±0.04) > MedGo-CoT (4.58±0.09) > DeepSeek-CoT (4.41±0.11) > MedGo (4.25±0.16) > DeepSeek (3.98±0.20) (Friedman test, p<0.001). In base mode, MedGo significantly outperformed DeepSeek (Holm-adjusted p=0.040). CoT improved both models (Holm-adjusted p<0.001 for DeepSeek; p=0.024 for MedGo) and reduced score dispersion. MedGo-CoT significantly outperformed DeepSeek-CoT in individualized adjustment (adjusted p<0.001) and dosing precision (adjusted p=0.005). ChatGPT–expert correlation was negligible (overall Kendall τ=0.153, p=0.003; subgroup τ=0.06–0.20, all p>0.05).

**Conclusions:** Domain-specific medical LLMs enhanced by CoT approach the antibiotic decision-making level of real physicians, with advantages in individualization and dosing precision. However, notable deficiencies persist in antimicrobial stewardship ecological awareness and automated evaluation reliability, underscoring the continued indispensability of senior clinical expertise.

## INTRODUCTION

Artificial intelligence (AI) is reshaping healthcare at an unprecedented pace. Transformer-based large language models (LLMs) have demonstrated near-human cognitive capabilities in complex reasoning, knowledge retrieval, and clinical text generation [1,2]. Contemporary LLMs offer a fundamentally new technological paradigm for clinical decision support [3].

Antimicrobial resistance (AMR) is one of the most urgent global health crises of the 21st century [4,5]. The core objective of antimicrobial stewardship (AMS) is to ensure “the right antibiotic, at the right dose, for the right duration”—optimizing therapeutic outcomes while minimizing selective pressure for bacterial resistance [6]. However, microbiological testing typically requires 2–5 days, and significant inter-patient variability in age, weight, hepatic and renal function, and comorbidities renders “one-size-fits-all” guidelines insufficient for truly personalized antimicrobial recommendations [7,8].

AI offers novel pathways to address these challenges. Machine learning models have demonstrated practical value in predicting pathogen susceptibility [9] and providing individualized antibiotic recommendations [10], achieving accuracy comparable to specialist physicians in complex diagnostic challenges [11]. Recent studies indicate that chain-of-thought (CoT) and other prompt engineering techniques can improve LLM accuracy in medical question answering and real-world case diagnosis [12,13]. Nevertheless, in infection control and antimicrobial stewardship, existing research has largely focused on monitoring prescribing behavior or training traditional machine learning methods from structured data [7,14], with less attention to LLM-driven clinical reasoning. The rapid evolution of LLMs has raised several critical questions: Can general-purpose LLMs handle the complexity of antibiotic prescribing decisions? Do domain-specific models—augmented with retrieval-augmented generation (RAG) and clinical reinforcement learning fine-tuning—offer substantive advantages? Can structured reasoning techniques such as CoT prompting improve prescribing quality [15]?

To address these questions, the present study employed a double-blind evaluation with a 2×2 factorial design to systematically compare a domain-specific medical LLM (MedGo) and a general-purpose LLM (DeepSeek V3.5) under both standard direct prompting and CoT prompting conditions, using real physician prescriptions as a clinical baseline. We further assessed the feasibility of using an LLM (ChatGPT 5.2) as an automated evaluation tool for antibiotic regimens.

## METHODS

### Study design

This retrospective, double-blind, randomized-sequence evaluation employed a 2×2 factorial design (model type: MedGo vs. DeepSeek; prompting strategy: standard direct prompt vs. CoT prompt) with an additional real clinical baseline control. For each of the 59 cases, five parallel regimens were generated: four from AI conditions (MedGo-Standard, MedGo-CoT, DeepSeek-Standard, DeepSeek-CoT) and one extracted from the medical record representing the original attending physician’s prescription. All regimens were anonymized through standardized formatting and independently evaluated by experts under blinded conditions.

### Case source and selection

The study consecutively enrolled patients receiving antibiotic therapy at the Intensive Care Unit (ICU) of Shanghai East Hospital between July 2025 and December 2025. Inclusion criteria: (1) age ≥18 years; (2) clinically diagnosed infection requiring antibiotic therapy, irrespective of the availability of microbiological or antimicrobial susceptibility testing (AST) results, or perioperative prophylactic antibiotic use; (3) complete electronic medical records covering demographic information, infection site, microbiological results, organ function indicators (eGFR, ALT/AST), allergy history, and the actual antibiotic regimen administered. Exclusion criteria: (1) missing critical decision-making data; (2) death within 24 h of admission, discharge against medical advice, or transfer to another hospital.

### AI agent configuration

#### Model architecture

MedGo (domain-specific medical LLM) uses a hybrid architecture combining a base model with retrieval-augmented generation (RAG) and fine-tuning. Through RAG, the model accesses a large, dynamically updated structured medical knowledge base comprising authoritative guidelines, drug package inserts, and recent literature, mitigating hallucination while preserving traceability. During alignment, MedGo underwent reinforcement learning from human feedback (RLHF) and deep fine-tuning on Chinese clinical anti-infective practice standards and high-quality medical evaluation datasets.

DeepSeek (general-purpose LLM) was accessed through the DeepSeek V3.5 API (deepseek-chat) and served as a general-purpose performance baseline.

#### Data structuring and input

Raw electronic medical record information was extracted and uniformly converted into anonymized, standardized JSON text covering five domains: (A) Patient characteristics; (B) Laboratory and microbiological indicators (PCT, CRP, WBC, smear/culture/susceptibility results, mNGS reports); (C) Host organ function (serum creatinine/eGFR, ALT/AST/bilirubin, INR/APTT, QTc interval); (D) Treatment context (current and prior antibiotic use, invasive procedures); (E) Local antibiotic formulary and departmental resistance patterns over the preceding 12 months. The complete JSON template is provided in Supplementary Material S1. The actual antibiotic regimen prescribed by the attending physician was excluded from the model input.

#### Prompting strategies

Both models were evaluated under two standardized prompting strategies. Complete prompt texts are detailed in Supplementary Materials S2 and S3.

Strategy 1—Standard Prompt (Zero-shot): The model assumed the role of a senior clinical pharmacist and was instructed to directly output antibiotic decisions with justifications. The prompt paradigm was: “You are a clinical pharmacist and a senior physician specializing in antibiotic use. I need you to make medication decisions… Please think carefully and answer briefly: 1. Which drug(s) to use, the dosage, and duration; 2. What is your rationale.”

Strategy 2—Chain-of-Thought (CoT) Prompt: The model was required to demonstrate explicit multi-stage clinical reasoning before generating the final prescription, following a four-step process: (1) Drug screening and matching; (2) Personalized dosing calculation and optimization (Cockcroft-Gault formula, hepatic adjustment, special populations); (3) Multi-dimensional ranking and regimen generation (efficacy, safety, AMS principles); (4) Explanatory output with guideline evidence, rationale, dose calculations, and monitoring recommendations.

#### Post-processing and blinding

To ensure double-blind integrity, all AI-generated and physician-written regimens underwent rigorous post-generation standardization. An automated script stripped characteristic formatting features, retaining only: antibiotic name, single dose, dosing frequency, route, duration, and a distilled core justification (maximum three sentences). The five regimens per case were randomly assigned de-identified labels (A/B/C/D/E).

Three senior clinicians (deputy chief physician of infectious diseases, chief physician of critical care medicine, and senior clinical pharmacist; each with >10 years of experience, none involved in model development) independently completed two rounds of scoring— comprehensive clinical rationality (1–5 scale) and five-domain sub-scores—using electronic forms without cross-discussion. A third-party research assistant performed unblinding and data compilation.

### Evaluation metrics

Primary outcome: comprehensive clinical rationality score (1–5 scale; 1=very poor, 5=excellent). Secondary outcomes: domain-specific sub-scores across five dimensions—(1) Antimicrobial spectrum coverage (weight 35%); (2) Dosing regimen precision (weight 25%); (3) Individualized adjustment (weight 15%); (4) Guideline concordance (weight 15%); (5) Pharmacoeconomic consideration (weight 10%). Complete definitions are provided in Supplementary Material S5.

### LLM-based automated evaluation

ChatGPT 5.2 was introduced as an independent automated evaluation tool, scoring each regimen on a 0–100 scale (antimicrobial spectrum coverage: 35 points; dosing regimen precision: 25; individualized adjustment: 15; guideline concordance: 15; pharmacoeconomics: 10). This metric was used solely to evaluate correlation with human expert scores via Kendall’s τ; it did not inform primary conclusions. Complete scoring guidelines are in Supplementary Material S6.

### Statistical analysis

Analyses were performed with Python 3.13 (SciPy 1.15, Pandas 3.0, NumPy 2.4). Continuous variables are reported as mean±SD or median. Inter-rater agreement was assessed with Kendall’s W. Overall group differences used the Friedman test, with pairwise post-hoc comparisons via Wilcoxon signed-rank test. P-values were adjusted with Holm-Bonferroni correction. Wilcoxon effect size r 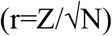is reported. Domain-level sub-score comparisons also used the Wilcoxon signed-rank test. ChatGPT–expert correlation was assessed with Kendall’s τ. Significance was set at p<0.05, and p<0.01 was considered highly significant.

## RESULTS

### Baseline characteristics

The study included 59 antibiotic-treated cases from the ICU of Shanghai East Hospital (age 70.6±15.2 years, range 25–95 years; 66.1% male). Antibiotic use scenarios, renal function distribution, and comorbidities are detailed in Table 1. The case mix was highly heterogeneous, covering most common ICU clinical scenarios with substantial complexity and genuine decision difficulty.

**Table 1.** Baseline characteristics of enrolled patients. Data are presented as mean±SD, n (%), or median (range). eGFR, estimated glomerular filtration rate.

| Characteristic | Value |
| --- | --- |
| Total cases | 59 |
| Age (years, mean±SD) | 70.6±15.2 |
| Male, n (%) | 39 (66.1%) |
| Infection type, n (%) |  |
| Perioperative prophylaxis | 20 (33.9%) |
| Mixed infections | 10 (16.9%) |
| Pneumonia | 9 (15.3%) |
| Other (urinary tract, etc.) | 5 (8.5%) |
| Intra-abdominal infections | 4 (6.8%) |
| Bloodstream infections | 4 (6.8%) |
| Skin and soft tissue infections | 3 (5.1%) |
| Other | 4 (6.8%) |
| Renal function |  |
| eGFR <60 mL/min/1.73 m<sup>2</sup> | 21 (35.6%) |
| eGFR ≥60 mL/min/1.73 m<sup>2</sup> | 38 (64.4%) |
| Comorbidities (mean±SD) | 7.0±4.1 |
| Range (median) | 1–19 (7) |
| >10 comorbidities, n (%) | 12 (20.3%) |

### Inter-rater agreement

Kendall’s W among the three experts was 0.52 (Friedman χ^2^=5.20, p=0.074), reflecting moderate agreement. With only five rated items, this represents an acceptable consistency level.

### Comprehensive score comparison

Comprehensive scores ranked as: real physicians (4.76±0.04) > MedGo-CoT (4.58±0.09) > DeepSeek-CoT (4.41±0.11) > MedGo (4.25±0.16) > DeepSeek (3.98±0.20). The Friedman test showed highly significant differences (χ^2^=34.82, p<0.001) (Table 2, Figure 1).

**Table 2.** Comprehensive scores across regimens (1–5 scale) Scores on a 1–5 scale (1=very poor, 5=excellent). Friedman test: χ^2^=34.82, p<0.001. CoT, chain-of-thought.

| Regimen | Expert 1 | Expert 2 | Expert 3 | Mean±SD |
| --- | --- | --- | --- | --- |
| DeepSeek (Standard) | 4.13 | 4.06 | 3.75 | 3.98±0.20 |
| DeepSeek-CoT | 4.33 | 4.53 | 4.37 | 4.41±0.11 |
| MedGo (Standard) | 4.42 | 4.22 | 4.11 | 4.25±0.16 |
| MedGo-CoT | 4.66 | 4.59 | 4.49 | 4.58±0.09 |
| Real Physicians | 4.80 | 4.76 | 4.72 | 4.76±0.04 |

**Figure 1.**
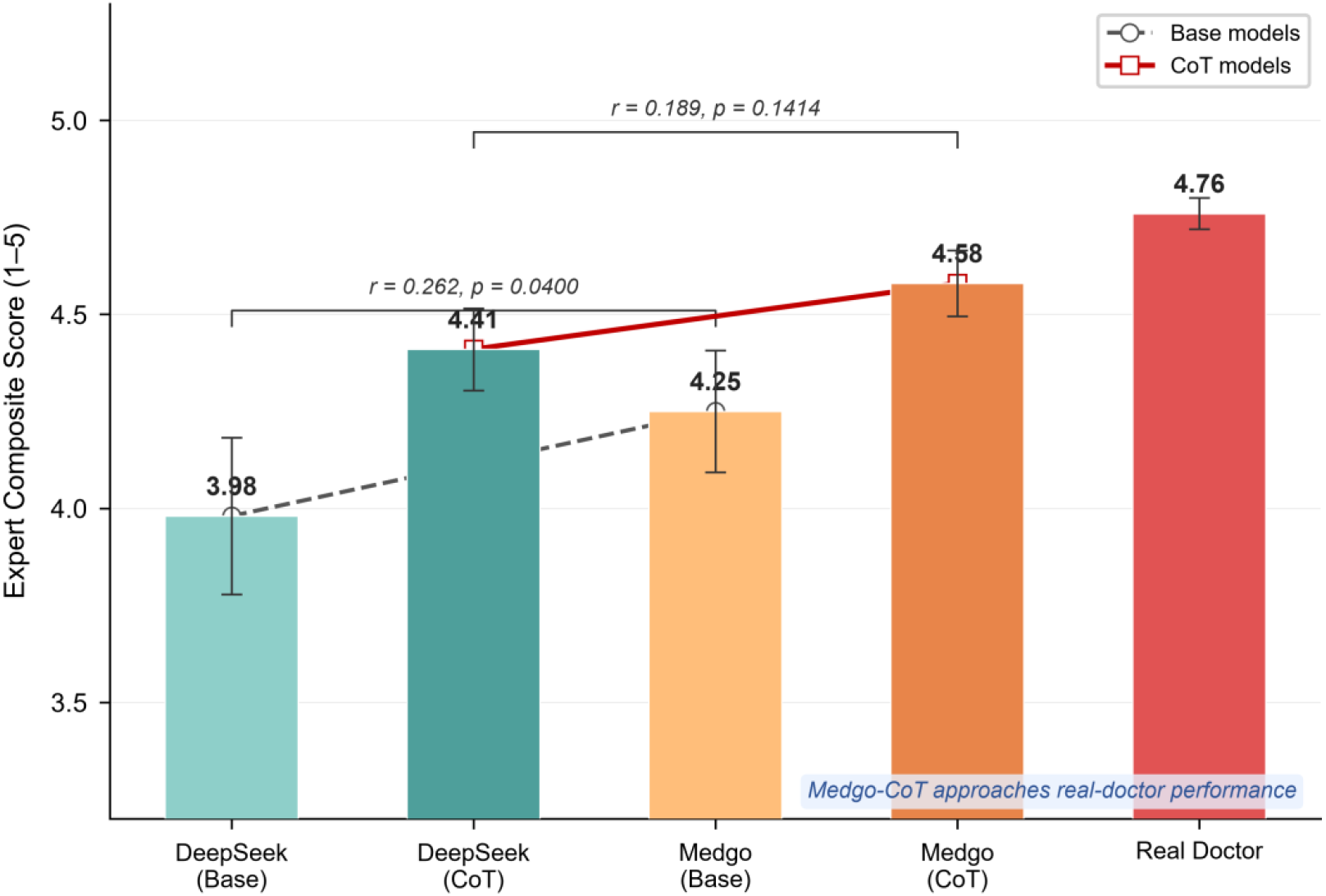
Comparison of comprehensive antibiotic prescription scores across models. Bar chart showing mean expert scores ± SD (1–5 scale). Dashed lines connect base models; solid lines connect CoT-enhanced models and real physicians. Friedman test: χ^2^=34.82, p<0.001. Holm-Bonferroni correction: MedGo significantly outperformed DeepSeek (adjusted p=0.040); CoT gains for both models were significant (both adjusted p<0.05); MedGo-CoT vs. real physicians was not significant (adjusted p=0.095).

In the base mode, MedGo significantly outperformed DeepSeek (Holm-Bonferroni adjusted p=0.040), demonstrating that the domain-specific medical LLM surpasses the general-purpose model at baseline. Under CoT enhancement, MedGo-CoT and DeepSeek-CoT showed no significant difference (adjusted p=0.141). The gap between MedGo-CoT and real physicians (0.18 points) was not statistically significant (adjusted p=0.095).

### CoT gain effect

CoT improved both models: DeepSeek from 3.98 to 4.41 (+0.43, adjusted p<0.001; W=80, r=0.485); MedGo from 4.25 to 4.58 (+0.33, adjusted p=0.024; W=99, r=0.351).

Both gains showed medium-to-large effect sizes. CoT reduced score dispersion: DeepSeek SD from 0.82 to 0.72; MedGo SD from 0.76 to 0.65 (Figures 2, 3). The majority of cases improved after CoT enhancement, with MedGo showing more uniform gains.

**Figure 2.**
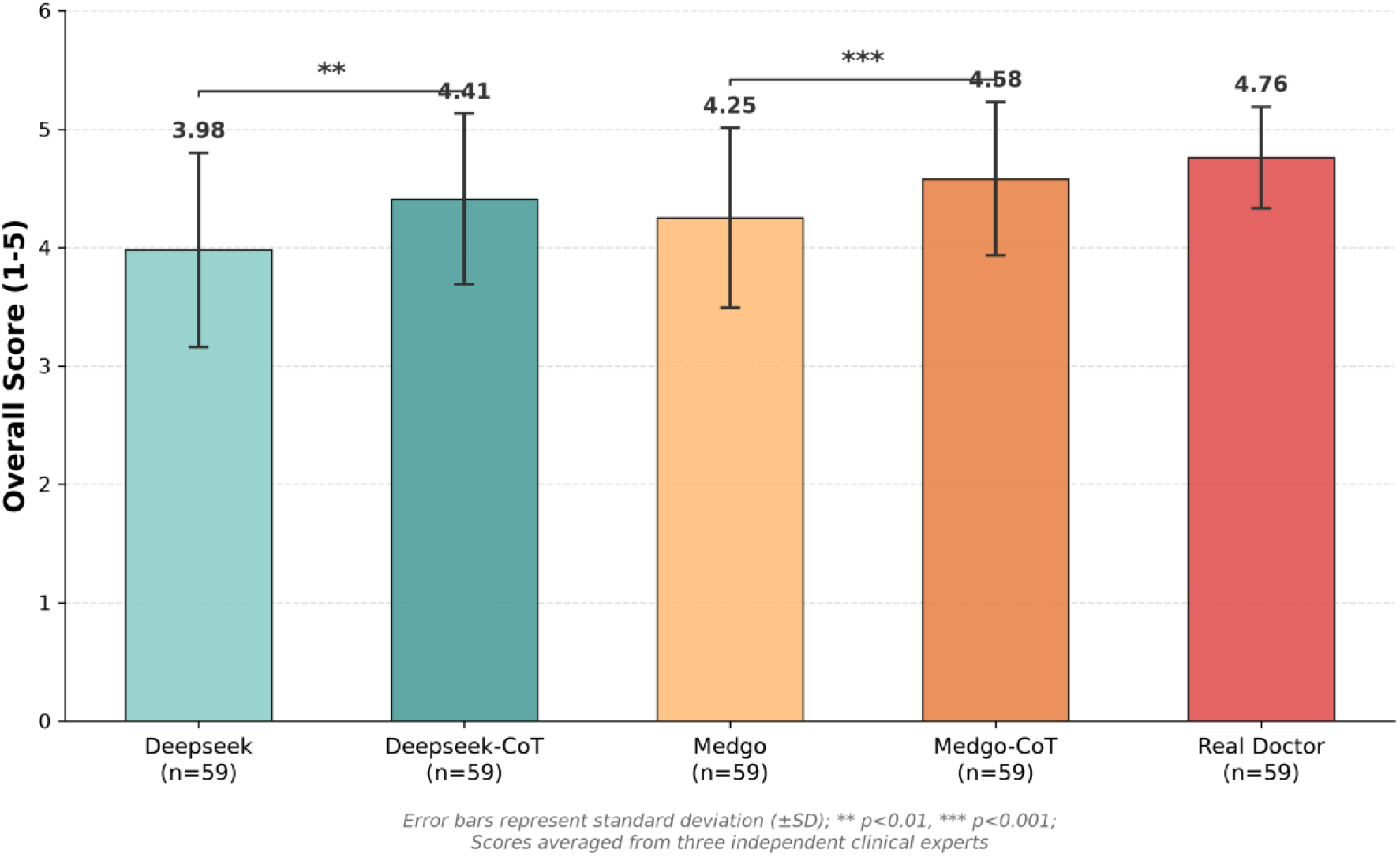
Gain effect and stability analysis of CoT prompting. Paired comparison of expert scores for DeepSeek and MedGo under standard vs. CoT prompting. DeepSeek: 3.98±0.82 → 4.41±0.72 (adjusted p<0.001); MedGo: 4.25±0.76 → 4.58±0.65 (adjusted p=0.024). Both r>0.4. CoT also reduced standard deviations.

**Figure 3.**
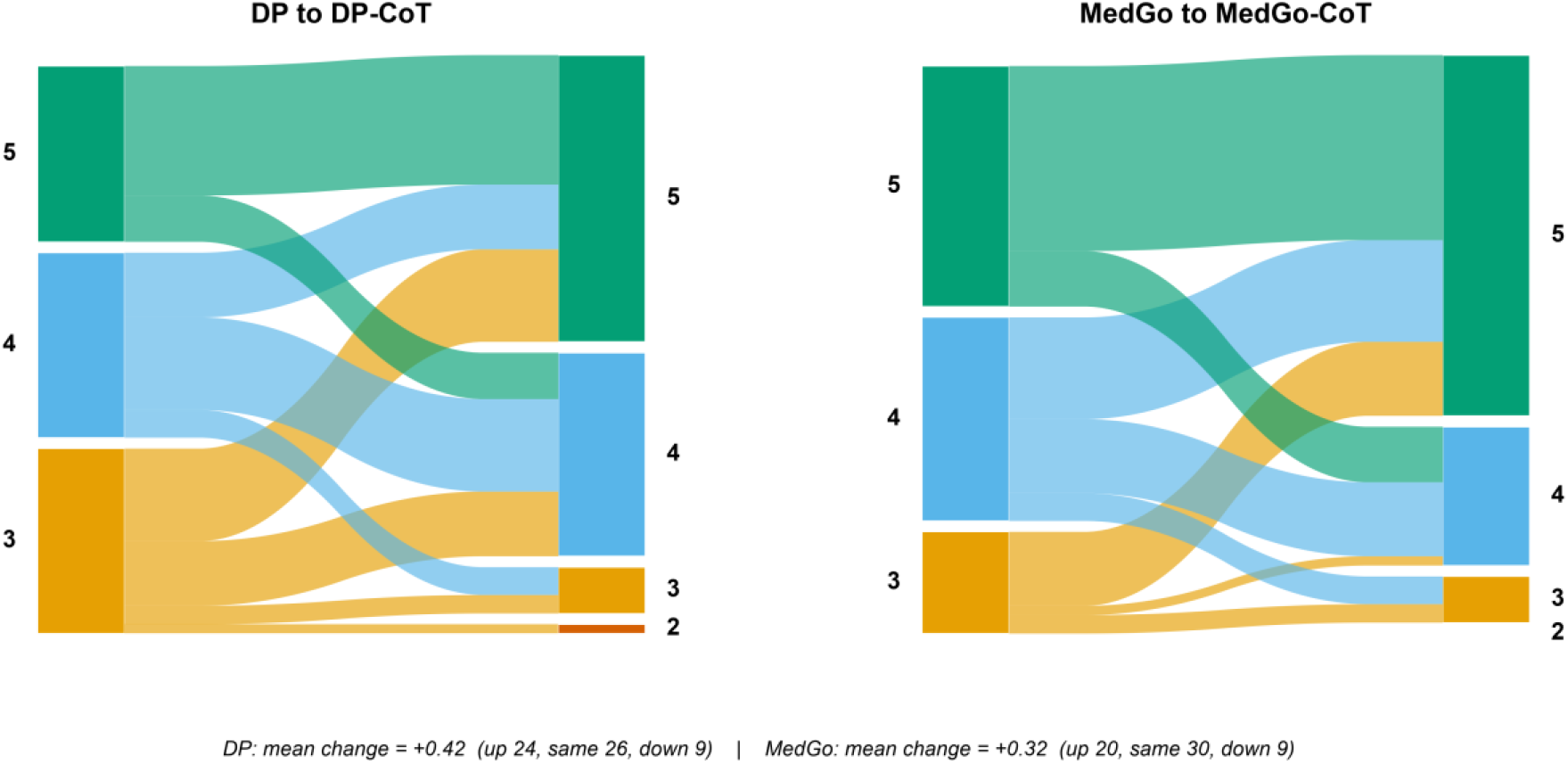
Sankey diagram of score transitions under CoT prompting. Left nodes: base version score ranges; right nodes: CoT-enhanced version. MedGo showed more uniform improvement.

### Multi-dimensional quality analysis

Dimension-level scores are in Table 3 and Figure 4. MedGo-CoT achieved the highest total score (4.08±0.47), significantly exceeding its base version (3.59±0.61, adjusted p<0.001) and numerically exceeding DeepSeek-CoT (3.87±0.46, adjusted p=0.130).

**Figure 4.**
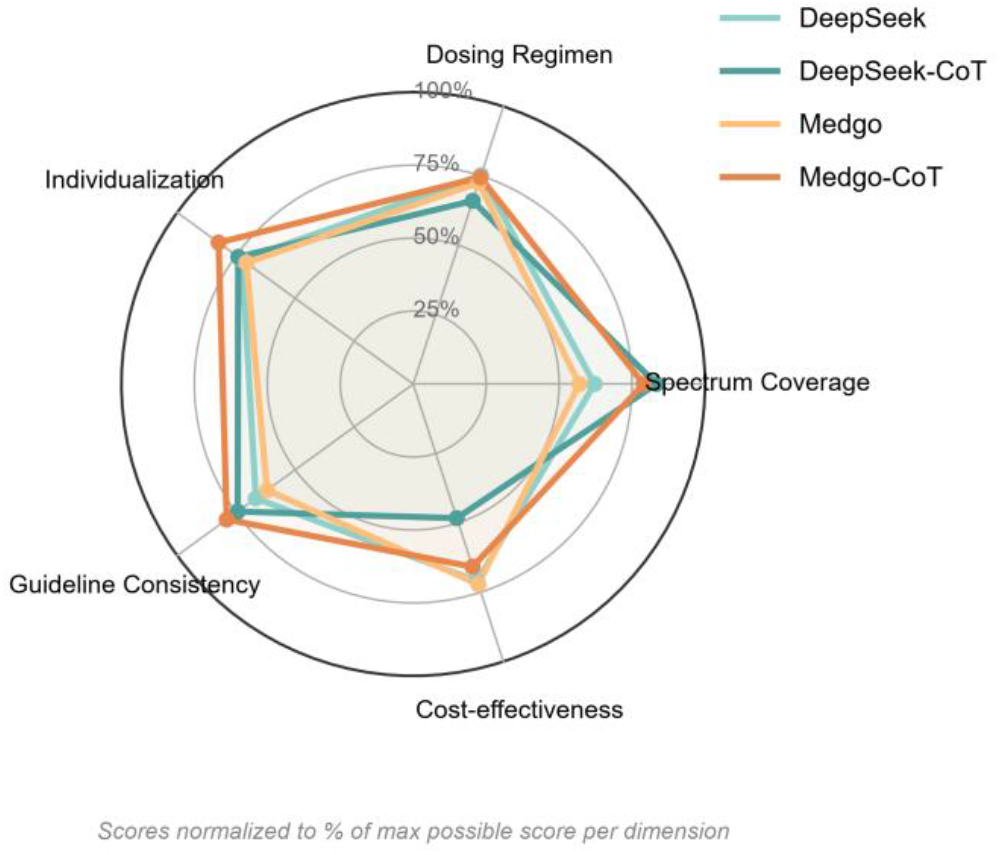
Radar chart of four AI conditions across five quality dimensions (1–5 scale). MedGo-CoT significantly outperformed DeepSeek-CoT in individualized adjustment (4.19±0.50) and dosing regimen precision (3.91±0.61) (both adjusted p<0.01). DeepSeek-CoT scored highest in antimicrobial spectrum coverage (4.38±0.85) but lowest in pharmacoeconomics (2.86±0.66).

**Table 3.** Dimension-level scores across models (1–5 scale) Æ,^b^,^c^ indicate MedGo-CoT significantly outperformed DeepSeek-CoT after Holm-Bonferroni correction (Æ p<0.001, ^b^ p=0.005, ^c^ p<0.001). Values are mean±SD.

| Dimension | DeepSeek | DeepSeek-CoT | MedGo | MedGo-CoT |
| --- | --- | --- | --- | --- |
| Antimicrobial spectrum coverage | $3.4 \pm 1.0$ | $4.4 \pm 0.9$ | $3.2 \pm 1.2$ | $4.1 \pm 0.9$ |
| Dosing regimen precision | $3.9 \pm 0.7$ | $3.6 \pm 0.6$ | $3.8 \pm 0.9$ | $3.9 \pm 0.6^b$ |
| Individualized adjustment | $3.8 \pm 0.6$ | $3.9 \pm 0.6$ | $3.8 \pm 0.5$ | $4.2 \pm 0.5^{\mathcal{A}}$ |
| Guideline concordance | $3.5 \pm 0.8$ | $3.9 \pm 0.6$ | $3.4 \pm 0.8$ | $4.1 \pm 0.6$ |
| Pharmacoeconomics | $3.8 \pm 0.8$ | $2.9 \pm 0.7$ | $3.9 \pm 0.6$ | $3.6 \pm 0.6^c$ |
| Total score | $3.7 \pm 0.7$ | $3.9 \pm 0.5$ | $3.6 \pm 0.6$ | $4.1 \pm 0.5$ |

MedGo-CoT showed its greatest advantages in individualized adjustment and dosing regimen precision. DeepSeek-CoT’s dosing precision score declined relative to its base version, suggesting a risk of “over-complication” with extended reasoning chains. Pharmacoeconomic scores declined in both CoT versions, possibly because the CoT emphasis on “full coverage” favored broader-spectrum, costlier antibiotics.

### ChatGPT automated evaluation vs. expert scores

Across 236 regimens, the overall Kendall τ between ChatGPT 5.2 and expert scores was 0.153 (p=0.003), with a negligible effect size (τ^2^≈0.023). Within individual model subsets, ChatGPT–expert correlations did not reach statistical significance (all p>0.05). The weak aggregate significance likely reflects inflated power from pooling rather than genuine evaluative validity (Figure 5).

**Figure 5.**
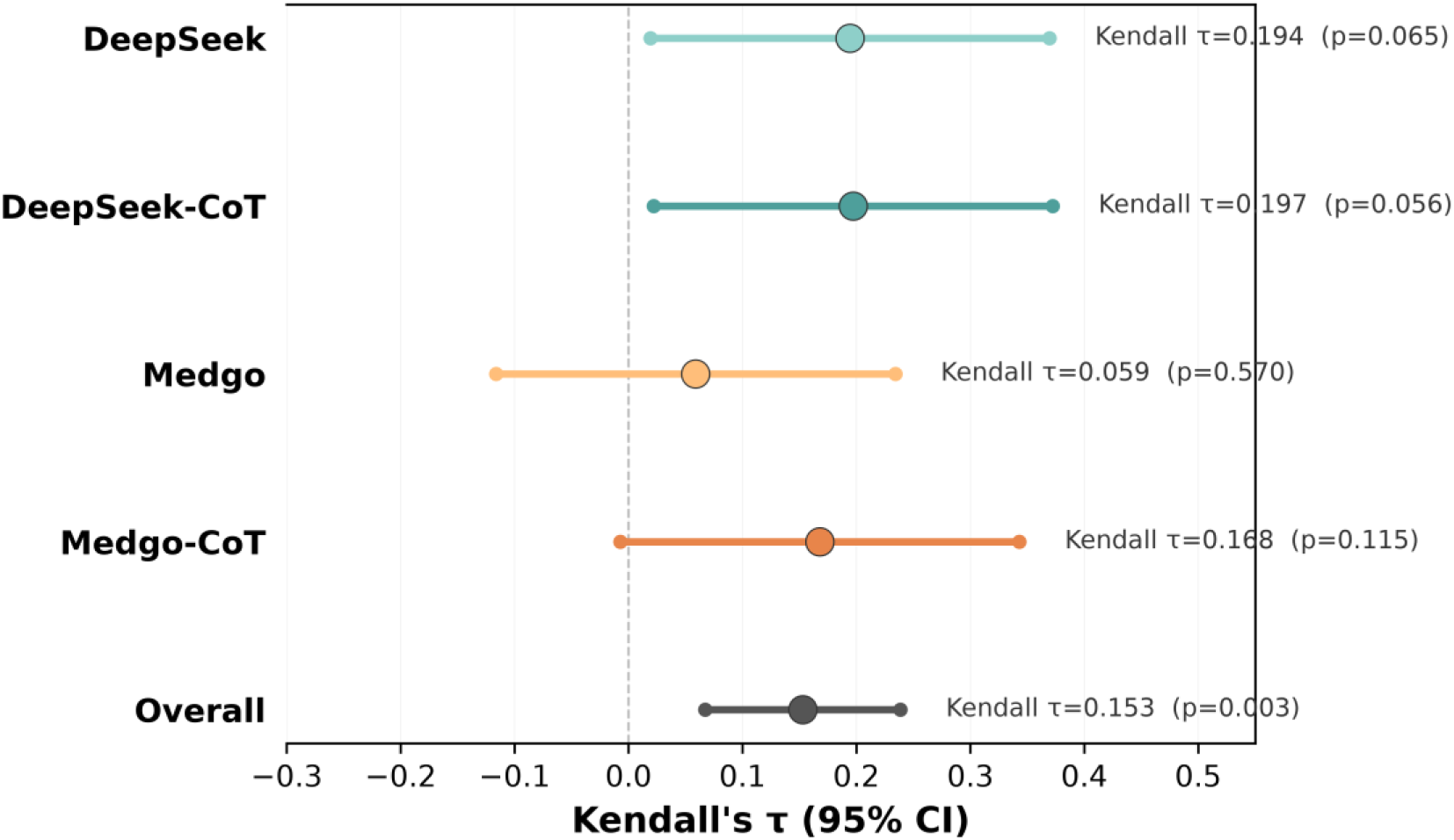
ChatGPT–expert score concordance (Kendall τ forest plot). Overall τ=0.153 (p=0.003; τ^2^≈0.023). No individual model reached significance (all p>0.05); significance emerged only after pooling. Error bars: 95% CI; dashed line: null (τ=0).

At the macro level, ChatGPT’s model ranking (MedGo-CoT 76.7 > DeepSeek-CoT 71.8 > DeepSeek 68.0 > MedGo 64.6) superficially aligned with expert trends. However, given the absence of individual-level correlation, this convergence does not support scoring accuracy.

## DISCUSSION

Through a double-blind systematic evaluation of 59 real-world complex infection cases, this study compared general-purpose and domain-specific LLMs in antibiotic prescribing decisions.

### Domain-specific medical large models enhance foundational medical capabilities

In the base mode, MedGo significantly outperformed DeepSeek. This advantage likely stems from MedGo’s domain knowledge infusion: pre-training and fine-tuning enforced medical safety boundaries, while the RAG-connected structured medical knowledge base enhanced the traceability of decision evidence. This domain specialization enabled MedGo to substantially reduce marginally acceptable (score 3) low-quality regimens, consistent with the view that domain specialization is key to medical AI [16,17].

### Chain-of-thought improves reasoning ability

Clinical antibiotic decision-making is inherently multi-step: from pathogen prediction and spectrum matching, to individualized dose adjustment, to AMS-based optimization. CoT prompting guides LLMs to decompose this process into traceable reasoning steps, reducing error accumulation from reasoning leaps. This is equivalent to introducing “intermediate computational scratchpads” within the autoregressive generation process, enabling the model to maintain intermediate reasoning states before final output [15].

Our results confirm this efficacy: DeepSeek improved by 0.43 and MedGo by 0.33, both with medium-to-large effect sizes. In individualized adjustment and dosing precision, MedGo-CoT showed the greatest advantages, while DeepSeek-CoT’s dosing score declined relative to baseline, suggesting over-complication risk with extended reasoning chains.

CoT also reduced score dispersion (DeepSeek SD: 0.82→0.72; MedGo SD: 0.76→0.65), enhancing output stability alongside quality. In clinical settings, decision stability itself is an important quality guarantee.

However, pharmacoeconomic scores declined in both CoT versions, likely because the emphasis on “full coverage” favored broader-spectrum, costlier antibiotics. This suggests that cost-effectiveness and AMS constraints must be explicitly incorporated into CoT prompt design, rather than treated as implicit.

### Persistent gaps in AI performance

Despite the significant improvements brought by domain-specific LLMs and CoT reasoning, systematic gaps remain in current AI for clinical application:

#### Insufficient clinical safety judgment and risk trade-off ability

Even with CoT-enhanced MedGo-CoT, a gap of 0.18 points from real physicians persisted. AI safety remains inadequate for elderly patients with fragile organ function.

For instance, for a 90-year-old patient with seemingly normal renal function, AI may still aggressively recommend a combination of “vancomycin + amikacin” in pursuit of “absolute coverage,” despite the high cumulative nephrotoxicity risk. This suggests that current AI follows rigid guideline dogmas and lacks the human capacity to “choose the lesser of two evils” [18,19].

#### Limited clinical situation recognition

AI has systematic difficulty distinguishing clinical contexts. First, it blurs prophylactic and therapeutic antibiotic use, treating perioperative cephalosporin prophylaxis as established infection. Second, it neglects source control—localized infections (e.g., confined intra-abdominal infection, acute cholecystitis) that could be managed with surgical drainage are instead escalated to higher-level antibiotics, reflecting a strong “fear of omission” [20]. Third, it exhibits mechanical spectrum stacking, such as prescribing “meropenem + metronidazole” while ignoring meropenem’s intrinsic anti-anaerobic activity.

#### Lack of antimicrobial stewardship ecological awareness

Anti-infective therapy affects not only the individual patient but the broader resistance ecology of the ward [21]. AI regimens showed three notable deficits: (1) Prioritizing efficacy over ecology—in long-stay ICU patients, AI favors “Gram-negative + Gram-positive” broad coverage to ensure “early effectiveness,” significantly increasing resistance selection pressure. (2) Lacking novel-agent stewardship—when meropenem and newer ceftazidime/avibactam (CAZ-AVI) are both viable, AI defaults to CAZ-AVI over concerns about carbapenem-resistant Klebsiella pneumoniae (CRKP) trends, disregarding the core AMS principle of preserving new drug durability. (3) Overdiagnosis of colonization and unnecessary escalation—AI misinterprets airway yeast colonization as invasive fungal infection, recommending costly second-line agents like isavuconazole, and blindly escalates from prolonged carbapenem use to polymyxins or tigecycline without susceptibility evidence or signs of infection progression, violating standard de-escalation/escalation logic.

#### Lack of tacit knowledge

More subtly, AI cannot capture the unwritten “tacit knowledge” in clinical practice [22]. For example, AI fails to understand why clinicians routinely administer antibiotics (e.g., vancomycin) for several days after ECMO decannulation to prevent catheter-related bloodstream infection—a common practice in many centers [23].

In summary, although MedGo-CoT can produce standardized evidence-based anti-infective “draft regimens,” it remains a system focused more on “the disease” than “the patient,” lacking the integrated clinical perspective of senior physicians and falling short of the requirements for AI in hospital infection prevention [24]. Human expert oversight—especially prescription authority and final verification for high-level antibiotics—remains indispensable under the current technological paradigm.

### LLM as an evaluation tool: a cautious positioning

ChatGPT 5.2 showed negligible correlation with expert scores, with complete loss of statistical significance within individual model subsets. The effect size (τ^2^≈0.023) indicates ChatGPT cannot explain more than 97.7% of expert rating variance. This finding accords with recent evaluations in antibiotic information provision [25] and stroke care [26], confirming that current LLM-based automated evaluation cannot substitute for expert judgment in fine-grained quality assessment [27,28], particularly in the safety-critical domain of antibiotic prescribing.

## Limitations

This study has several limitations. First, its single-center retrospective design and modest sample size (59 cases) constrain generalizability. Second, the weighted scoring framework may not capture all clinically relevant dimensions. Third, all evaluated models are time-bound; more recent models may yield different results. Fourth, generalizability to non-antibiotic medical domains requires further validation.

## CONCLUSION

This double-blind, 2×2 factorial evaluation demonstrates that domain-specific medical LLMs enhanced by CoT prompting (MedGo-CoT) achieve strong performance in antibiotic prescribing decisions. CoT prompting improves both the quality and consistency of LLM-generated prescriptions. However, current LLMs remain deficient in grasping clinical prescribing contexts, AMS ecological management, and tacit knowledge, lacking the integrated clinical judgment and risk trade-off capacity of senior clinicians. Human expert oversight remains indispensable. Our findings provide evidence-based guidance for developing and deploying LLM-driven clinical decision support systems for antimicrobial stewardship and position LLM-as-a-Judge as an adjunctive, rather than substitutive, tool in anti-infective therapy evaluation.

## Data Availability

All data produced in the present study are available upon reasonable request to the authors

## DECLARATIONS

### Competing interests

All authors have completed the ICMJE uniform disclosure form and declare: no support from any organisation for the submitted work; no financial relationships with any organisations that might have an interest in the submitted work in the previous three years; no other relationships or activities that could appear to have influenced the submitted work.

### Funding

This research received no specific grant from any funding agency in the public, commercial, or not-for-profit sectors.

## Acknowledgments

None.

## Contributorship

YL and HZ conceived and designed the study. CZ, FW, and WX collected and organized the clinical data. YZ and SM performed the statistical analysis. YL drafted the manuscript. All authors reviewed and approved the final version. HZ is the guarantor.

## Ethics approval

This study was approved by the Ethics Committee of Shanghai East Hospital, Tongji University School of Medicine. Given the retrospective design using de-identified data, the requirement for informed consent was waived.

## Notes

### Competing Interest Statement

The authors have declared no competing interest.

### Author Declarations

This study was approved by the Ethics Committee of Shanghai East Hospital, Tongji University School of Medicine.

