## supplemental File for "General-Purpose vs. Domain-Specific Large Language Models in Antibiotic Clinical Decision-Making: A Double-Blind Evaluation with a 2×2 Factorial Design"

### Supplementary Material: Detailed Methods

#### S1. Standardized Data Input Format (JSON Template)

The JSON template below defines all data fields extracted from electronic medical records for each case. Raw EMR data were converted into this standardized, de-identified format. Fields cover: (A) patient demographics and clinical characteristics, (B) laboratory and microbiological findings, (C) organ function status, (D) treatment background, and (E) local antibiotic formulary and resistance profiles. The actual treating physician's antibiotic regimen was strictly excluded from the input data fed to the models.

```
{
  "A_Patient_Characteristics": {
    "1_Demographics": {"Age": "", "Sex": "", "Weight": "", "Height": ""},
    "2_Clinical_Diagnosis": {"Infection_Site": "", "Severity_Score": "",
                            "Comorbidities": "", "Allergy_History": ""}
  },
  "B_Laboratory_and_Microbiology": {
    "1_Infection_Markers": {"PCT": "", "CRP": "", "WBC": "",
                           "Neutrophil_Percentage": "", "IL_6": "",
                           "PCT_Trend": "", "CRP_Trend": ""},
    "2_Microbiological_Evidence": {"Smear_Culture": "", "AST_Results": "",
           "Report_Time": "", "Specimen_Type": "", "Collection_Time": "",
           "Gram_Stain": "", "Pathogen_Name": "", "Resistance_Genes": ""},
    "3_mNGS_Report": ""
  },
  "C_Organ_Function": {
    "1_Renal": {"Serum_Creatinine": "", "Baseline_Creatinine": "", "BUN": "",
               "Ccr": "", "eGFR": ""},
    "2_Hepatic": {"Total_Bilirubin": "", "Direct_Bilirubin": "", "ALT": "",
                  "AST": "", "ALP": "", "GGT": "", "Albumin": "", "PT": ""},
    "3_Coagulation": {"INR": "", "APTT": "", "Platelet_Count": ""},
    "4_Other": {"QTc_Interval": ""}
  },
  "D_Treatment_Background": {
    "1_Current_and_Prior_Antibiotics": {
        "Current_Antibiotic": "", "Current_Dose": "",
        "Start_Time": "", "Duration": "",
        "Antibiotics_Past_90_Days": "", "Past_Course_Details": ""},
    "2_Invasive_Procedures": {"CVC": "", "Urinary_Catheter": "",
        "Mechanical_Ventilation": "", "Surgery_Drainage": "",
        "Indwelling_Duration": ""}
  }
}
```

#### S2. Standard Prompt (Zero-Shot)

This is the exact standard prompt text provided to both MedGo and DeepSeek under the Standard Prompt condition. The model was assigned the role of senior clinical pharmacist and asked to directly output an antibiotic regimen with supporting rationale, limited to 500 Chinese characters.

**Prompt Text**

```
"You are a clinical pharmacist and senior physician in antibiotic
use. I need you to make a medication decision. Select from the
following list of available antibiotics and determine the antibiotic
regimen for this patient at the time of the last laboratory result.
Please think carefully and answer the following questions concisely,
within 500 characters:
1. Which drug or drug combination to use, the dosage, and the
   duration of treatment.
2. The rationale for your choice (considering the patient's age,
   hepatic and renal function, and any supporting clinical guidelines
   or drug labeling)."

"The following is the list of available antibiotics:
Penicillin G (Benzylpenicillin)
Amoxicillin (Oral)
Ceftobiprole Medocaril Sodium (Injection)
Cefeprole Hydrochloride (Injection)
Cefprozil (Dispersible Tablet)
Cefdinir (Dispersible Tablet)
Cefuroxime Sodium (Injection)
Cefaclor (Dry Suspension)
Cefcapene Pivoxil Hydrochloride (Granules)
Cefixime (Tablet)
Cefmetazole Sodium (Injection)
Cefoperazone Sodium/Sulbactam Sodium (Injection)
Ceftriaxone Sodium (Injection)
Cefotaxime Sodium (Injection)
Cefotaxime Sodium/Tazobactam Sodium (Injection)
Ceftazidime/Avibactam Sodium
Ceftazidime (Injection)
Cefditoren Pivoxil (Granules)
Cefoxitin Sodium (Injection)
Cefazolin Sodium (Injection)
Imipenem/Cilastatin
Meropenem
Aztreonam
Amikacin
Azithromycin
Tigecycline
Vancomycin
Levofloxacin
Moxifloxacin
Trimethoprim-Sulfamethoxazole (SMZ-TMP)
Clindamycin
Metronidazole
Ornidazole
Rifampin
Linezolid
Fosfomycin Sodium
Polymyxin B
Polymyxin E (Colistin)"
```

#### S3. Chain-of-Thought (CoT) Prompt

This is the exact CoT prompt text provided to both MedGo and DeepSeek under the CoT condition. The prompt enforces a structured four-step clinical reasoning process, including explicit scoring weights, dosing calculation requirements, and a mandatory output format template.

**Prompt Text**

```
"# Role Definition
You are an experienced ICU clinical pharmacist and infectious
disease specialist, expert in antimicrobial treatment decisions
for critically ill patients. Your core task is: based on the
structured patient data provided, strictly follow the four-step
decision process below to generate an individualized, actionable,
well-supported antibiotic treatment plan.

# Input Data Description
You will receive patient information in the following categories
(some fields may be missing; make decisions based on available
data):
- A. Patient Characteristics: age, sex, weight, height, primary
  diagnosis, infection site, SOFA/APACHE II score, comorbidities,
  allergy history.
- B. Laboratory & Microbiology: PCT, CRP, WBC, smear/culture/
  susceptibility results (with sampling time and specimen source),
  mNGS report.
- C. Host Organ Function: serum creatinine, BUN, total bilirubin,
  ALT, AST, INR, ECG/QTc (if available).
- D. Treatment Background: current antibiotics (name, dose, start
  time, duration), antibiotics used in past 90 days, invasive
  procedures (mechanical ventilation, catheters, etc.).
- E. Local Knowledge: hospital antibiotic formulary, department-
  level antibiogram for the past 12 months (e.g., CRKP, MRSA,
  ESBL rates).

# Core Decision Process
Please strictly follow the four steps below and present the key
conclusions of each step in the final output.

## Step 1: Initial Drug Screening and Matching
1.1 Determine the treatment pathway:
    - If valid susceptibility data exist (sample collected within
      7 days and specimen source matches the current infection
      site) -> Targeted Therapy Pathway: select all "Sensitive (S)"
      and selected "Intermediate (I)" agents.
    - Otherwise -> Empiric Therapy Pathway: select empiric regimens
      from the knowledge base based on infection site, disease
      severity, and resistance risk factors.
1.2 Cross-reference with hospital formulary: retain only agents
    available at this institution. Warn if no suitable agent exists.

## Step 2: Individualized Dose Calculation and Optimization
For each drug from Step 1, perform the following:
2.1 Renal Adjustment:
    - Calculate creatinine clearance (Ccr) using the Cockcroft-Gault
      formula based on the most recent serum creatinine.
    - Adjust dose and dosing interval based on Ccr.
    - For narrow-therapeutic-index drugs (e.g., vancomycin,
      aminoglycosides), specify target TDM parameters (e.g., target
      peak concentration XX, trough concentration XX).
2.2 Hepatic Considerations:
    - Identify drugs primarily metabolized by the liver (e.g.,
      rifampin, azole antifungals).
    - Provide warnings or dose reduction recommendations based on
      bilirubin, ALT/AST levels.
2.3 Special Populations:
    - Elderly (≥65 years): consider reduced starting dose.
    - Obese (BMI ≥30): calculate dose based on adjusted body weight.
    - Pediatrics: calculate dose by weight (mg/kg) or body surface
      area.

## Step 3: Multi-Dimensional Ranking and Regimen Generation
Score all available agents from Step 2 based on the following
weighted priorities, output 1-3 preferred regimens with clear
ranking (first-line, second-line, alternative).

Scoring Weights (in descending order of priority):
1. Efficacy First:
   - Certainty of pathogen coverage (Targeted > Empiric)
   - Bactericidal > Bacteriostatic (especially for septic shock)
   - Local susceptibility data support (e.g., >90% susceptibility
     rate = full score)
2. Safety Optimization:
   - Match allergy history (absolute exclusion)
   - Match hepatic/renal function (exclude contraindicated agents)
   - Avoid agents with QTc prolongation risk
   - Assess drug-drug interactions with concurrent medications
3. Antimicrobial Stewardship (AMS) Principles:
   - Given equal efficacy: narrow-spectrum > broad-spectrum
   - Monotherapy > combination therapy (unless clear indication:
     septic shock, MDR organisms, infective endocarditis, etc.)
   - Cost-effectiveness: when efficacy and safety are comparable,
     select the more economical option

## Step 4: Generate Explanatory Output
For each preferred regimen (especially first-line), generate the
structured explanation below. This is key to clinician trust in
the system.

### Output Format Template:

#### Regimen 1: [Drug Name] [Dose] [Frequency] [Route] (First-Line)

**Core Rationale:**
1. Guideline Match: [cite specific guideline name, year, section,
   and level of evidence]
2. Patient Match: [explain why this regimen suits the patient's
   infection site, severity, and comorbidities]
3. Local Data Support: [cite institutional antibiogram data,
   expected efficacy]
4. Susceptibility Support (if Targeted Therapy pathway): [cite
   specific AST results]

**Dose Calculation Details:**
- Patient weight: [XX] kg, Height: [XX] cm
- Creatinine clearance (Ccr): [XX] mL/min (Cockcroft-Gault formula)
- Dose adjustment rationale: [explain adjustment logic based on
  Ccr/hepatic function]

**Key Warnings and Recommendations:**
- Risk Warnings: [e.g., nephrotoxicity, hepatotoxicity, QTc
  prolongation, seizure risk, etc.]
- De-escalation/Switch Prompt: [suggest re-evaluation after 48-72
  hours, consider switching to XX if AST results become available]
- Monitoring Recommendations: [blood cultures, PCT, therapeutic
  drug monitoring, etc.]
- Duration Recommendation: [suggest total course of XX days, or
  adjust dynamically based on clinical response]

#### Regimen 2: [Drug Name] [Dose] ... (Second-Line)
[Briefly explain the key differences from the first-line regimen]

#### Regimen 3: [Drug Name] [Dose] ... (Alternative)
[Brief explanation]

---

### Missing Data Handling
- If critical data are missing (e.g., no recent creatinine, no
  weight), you must: (a) explicitly warn in the output, and (b)
  state the limitations of your recommendation.
- Warning format: "[!] Data Missing Warning: [specific missing
  item(s)]. This recommendation is based on [assumption/default].
  Please verify with clinical data."

### Final Output Requirements
- Follow the Output Format Template above.
- All doses, frequencies, and routes must be clear and unambiguous.
- Avoid vague terms such as "possibly" or "perhaps" unless in the
  context of uncertainty warnings.
- If no reasonable regimen can be generated, state the reason
  explicitly and recommend clinical consultation.

---

Select only from the following available antibiotics:
Penicillin G (Benzylpenicillin)
Amoxicillin (Oral)
Ceftobiprole Medocaril Sodium (Injection)
Cefeprole Hydrochloride (Injection)
Cefprozil (Dispersible Tablet)
Cefdinir (Dispersible Tablet)
Cefuroxime Sodium (Injection)
Cefaclor (Dry Suspension)
Cefcapene Pivoxil Hydrochloride (Granules)
Cefixime (Tablet)
Cefmetazole Sodium (Injection)
Cefoperazone Sodium/Sulbactam Sodium (Injection)
Ceftriaxone Sodium (Injection)
Cefotaxime Sodium (Injection)
Cefotaxime Sodium/Tazobactam Sodium (Injection)
Ceftazidime/Avibactam Sodium
Ceftazidime (Injection)
Cefditoren Pivoxil (Granules)
Cefoxitin Sodium (Injection)
Cefazolin Sodium (Injection)
Imipenem/Cilastatin
Meropenem
Aztreonam
Amikacin
Azithromycin
Tigecycline
Vancomycin
Levofloxacin
Moxifloxacin
Trimethoprim-Sulfamethoxazole (SMZ-TMP)
Clindamycin
Metronidazole
Ornidazole
Rifampin
Linezolid
Fosfomycin Sodium
Polymyxin B
Polymyxin E (Colistin)"
```

#### S4. Output Post-Processing and Blinding Protocol

To ensure the integrity of double-blind evaluation, all AI-generated and physician-extracted regimens underwent standardized post-generation formatting to remove stylistic markers that could reveal their origin (e.g., AI-typical disclaimers, markdown formatting, summary-conclusion structures).

Each regimen was manually reformatted to retain only: drug name(s), single dose, dosing frequency, route of administration, treatment duration, and a distilled core rationale (limited to 3 sentences). The five formatted regimens per case were randomly labeled A, B, C, D, and E before distribution to the expert panel. The five regimens comprised: four AI-generated (MedGo-Standard, MedGo-CoT, DeepSeek-Standard, DeepSeek-CoT) and one extracted from the actual treating physician's medical record.

Experts completed evaluations independently using electronic forms, with cross-discussion prohibited. Unblinding and data aggregation were performed by an independent research assistant after all evaluations were completed.

#### S5. Five Evaluation Dimensions — Full Definitions

The primary outcome was a Clinical Appropriateness Composite Score (1 item, 1-5 scale). Secondary outcomes comprised five dimension-specific scores (1 = very poor, 5 = excellent). The dimensions were designed to align with the ChatGPT automated scoring framework for cross-validation purposes.

**Dimension 1: Spectrum Coverage**

Assesses whether the selected antibiotic(s) accurately cover the target pathogen(s) and are concordant with antimicrobial susceptibility testing results.

**Dimension 2: Dosing Regiment**

Assesses whether the single dose, dosing frequency, route of administration, and total treatment duration conform to pharmacokinetic/pharmacodynamic (PK/PD) principles.

**Dimension 3: Individualization**

Assesses whether the regimen incorporates precise dose adjustments or special considerations based on the patient's specific pathophysiological state, including age, body weight, dynamic hepatic/renal impairment, and extracorporeal blood purification status.

**Dimension 4: Guideline Consistency**

Assesses whether the regimen strictly adheres to authoritative clinical guidelines and successfully avoids the patient's documented allergies, comorbidity contraindications, and serious drug-drug interactions.

**Dimension 5: Economic Rationality**

Assesses whether the regimen aligns with Antimicrobial Stewardship (AMS) economic principles, and whether there is evidence of unwarranted use of expensive broad-spectrum antibiotics or unnecessary consumption of healthcare resources.

#### S6. ChatGPT Automated Evaluation — Full Scoring Rubric and Prompt

To explore the feasibility of automated evaluation in medical LLM assessment, ChatGPT 5.2 was deployed as an independent automated judge. All formatted regimens and authoritative guideline texts were systematically input to the model, which was instructed to assign scores on a 0-100 scale using the following predefined rubric and evaluation prompt.

**Scoring System Overview (5 items, total 100 points)**

(1) Spectrum Coverage — maximum 35 points;

(2) Dosing Precision — maximum 25 points;

(3) Individualization — maximum 15 points;

(4) Guideline Consistency — maximum 15 points;

(5) Economic Rationality — maximum 10 points.

**Complete Evaluation Prompt**

The following is the exact prompt text provided to ChatGPT 5.2 for each evaluation task:

```
"You are a clinical infectious disease and clinical pharmacy
expert. Please score the first-line antibiotic regimens generated
by the four models strictly according to the fixed scoring rubric
below.

Scoring Objective: Ensure consistent scoring across repeated
evaluations for the same case. Do not assign an overall score
based on general impression; you must score each dimension
individually and then compute the total.

[Patient Data]
{case_text}

{model_blocks}

[Scoring Rubric (Total: 100 points)]

1. Spectrum Coverage (35 points)
   - Complete coverage of primary suspected/confirmed pathogen: 35
   - Basically adequate, minor coverage gaps: 25-30
   - Notable coverage deficiencies: 10-20
   - Key pathogen(s) not covered: 0-10

   Specific Deductions:
   - Misses coverage of confirmed resistant organisms (e.g., CRE,
     CRAB, MRSA): -10 per item
   - Lacks anti-pseudomonal coverage when indicated: -8

2. Dosing and Administration Regimen (25 points)
   - Dose, frequency, and infusion method fully appropriate: 25
   - Minor issues present: 18-22
   - Notable dosing errors or inappropriate infusion: 8-17
   - Severe errors: 0-7

   Specific Deductions:
   - No renal function adjustment: -8
   - CRRT dose error: -10
   - Extended/continuous infusion indicated but not used: -3

3. Individualization Considerations (15 points)
   Factors include: hepatic and renal function, body weight/BMI,
   allergy history, infection site penetration, prior antibiotic
   exposure history.
   - Fully considered: 15
   - Partially considered: 8-12
   - Notable omissions: 0-7

4. Guideline Consistency (15 points)
   References: IDSA guidelines, ESCMID guidelines, Sanford Guide,
   Chinese antimicrobial therapy guidelines.
   - Fully concordant: 15
   - Basically concordant: 10-13
   - Notable deviations: 0-9

5. Economic Rationality and Stewardship (10 points)
   - Preference for narrow-spectrum / reasonable de-escalation /
     appropriate cost: 8-10
   - Acceptable: 5-7
   - Notable overtreatment or economically unreasonable: 0-4

[Output Requirements]
You must list the score for each dimension separately, then
compute the total score.

Format:
1. Spectrum Coverage: X/35
2. Dosing Regimen: X/25
3. Individualization: X/15
4. Guideline Consistency: X/15
5. Economic Rationality: X/10
 Total Score: X/100
Deduction Rationale:
- xxx
- xxx


Rules:
- If first-line regimens from multiple models are essentially
  identical (same drug(s), dose, frequency, and combination),
  they must receive the same score (+/-1 point tolerance).
- Scoring must not be influenced by language expression quality.
- If information is insufficient, do not penalize a model for
  declining to guess."
```

These ChatGPT-generated scores were not used to determine the primary conclusions regarding model superiority or clinical utility. Their sole purpose was to enable Spearman rank correlation analysis for validating the concordance and robustness of automated AI evaluation relative to expert human assessment.
